# Effectiveness of mepolizumab in a real-life cohort of patients with severe refractory eosinophilic asthma and multiple comorbidities

**DOI:** 10.1101/2020.05.26.20112052

**Authors:** Claudia Crimi, Raffaele Campisi, Giulia Cacopardo, Rossella Intravaia, Santi Nolasco, Morena Porto, Corrado Pelaia, Nunzio Crimi

## Abstract

**BACKGROUND:** Patients with severe asthma often suffer from comorbidities whose impact on the course of biological therapy has not been elucidated yet.

**OBJECTIVE:** To evaluate real-life effectiveness and the presence/absence of predictors of treatment response in patients with one or more comorbidities who received mepolizumab (MEPO) for the treatment of severe eosinophilic asthma (EA).

**METHODS:** Health records of 31 patients were retrospectively analyzed. Asthma control test (ACT) score, blood eosinophil count, forced expiratory volume in 1 second (FEV_1_), FEV_1_% of predicted and FEV_1_/FVC (Forced Vital Capacity) ratio, oral corticosteroid (OCS) dosage and exacerbations were recorded at baseline (T0), after 3 (T1), 6 (T3), nine (T6) and 12 months (T12). A clinical response was defined as: i) 30% exacerbation decrease; ii) 80% blood eosinophilia reduction; iii) 3 point ACT increase; iv) FEV_1_ increase ≥ 200 mL.

**RESULTS:** At T12 blood eosinophil level decreased by 89.89% (p>0.0001), an improvement in ACT of 3 points from baseline was recorded in 80.65% of patients (p>0.0001) and 96.77% of patients reduced by minimum 30% the number of exacerbations (p>0.0001). 84% of patients discontinued OCS (p>0.0001). FEV_1_ increased by 0.22 (p=0.0224) while FEV_1_/FVC was statistically significant only at T1. No significant differences were generally found among patients with a specific comorbidity. The number of comorbidities did not influence treatment response. Neither the comorbidities nor other characteristics (sex, BMI, age, smoking, baseline eosinophil level) influenced treatment response.

**CONCLUSIONS:** MEPO in patients with severe EA is effective regardless of the presence of one or more comorbidities.

## 1. Introduction

Severe eosinophilic asthma is a subtype of asthma characterized by persistent eosinophilic airway inflammation and recurrent exacerbations despite treatment with high doses of glucocorticoids [1]. In the last decade, several biological molecules with a steroid-sparing effect have been introduced in the field of severe asthma. Mepolizumab (MEPO) is an IgG1/k class humanized monoclonal antibody approved in patients ≥12 years of age for the treatment of moderate-to-severe eosinophilic asthma owing to its ability to block circulating interleukin-5 (IL-5) responsible for eosinophils development, maturation and survival [2]. In large placebo-controlled trials, treatment with MEPO was well tolerated, resulting in a substantial fall in blood eosinophils and a significant reduction of intake/dosage of oral corticosteroids (OCS), reduction of exacerbations and an overall improvement of lung function [3-6]. In practice, MEPO was shown to change the course of severe eosinophilic asthma thanks to its ability to reduce asthma exacerbation rates and improve the quality of life in these patients, as clearly outlined in a meta-analysis of seven randomized controlled trials (RCTs) [7]. Moreover, severe eosinophilic asthma, just like asthma, can be associated with several comorbidities (e.g. nasal polyposis, gastro-esophageal reflux disease (GERD), bronchiectasis, allergic and nonallergic rhinitis, obesity) which have a consistent impact on treatment outcome, asthma symptoms, risk of exacerbations and patient’s quality of life [8-11]. Recently, researchers have been trying to identify, based on the presence of comorbidities or lifestyle habits (i.e. smoking), specific asthma phenotypes with the ultimate goal of personalizing the therapeutic approach. However, at present, the characteristics of these phenotypes and the impact of treatment on each of them are still not fully answered questions [12]. Thus, the monitoring of new biological agent effectiveness in real-life practice may provide, in a heterogeneous disease like asthma, relevant data complementary to those of randomized control trials [13]. Moreover, a detailed assessment of comorbidities in patients with severe eosinophilic asthma is important for clinical practice and, to the best of our knowledge, has not been outlined yet. Under this perspective, we retrospectively examined a group of patients with multiple comorbidities who received MEPO for the treatment of severe eosinophilic asthma in order to evaluate its real-life effectiveness and to explore the presence/absence of potential predictors for treatment response.

## 2. Methods

### 2.1. Study design and subjects

This was a single-centre, retrospective study based on health records of patients who consulted a specialist from January 2018 to June 2019 at the Respiratory Medicine Unit – A.O.U. “Policlinico-Vittorio Emanuele”, Catania – Italy. All outpatients ≥12 years of age prescribed with MEPO were included in the study. Severity at baseline was defined according to the GINA guidelines [14]. All patients met the criteria for severe uncontrolled asthma according to the ATS/ERS guidelines [1] and received MEPO 100 mg subcutaneously every 4 weeks from T0 for at least 12 months (T12). All patients had >150 eosinophils/μl and a history of at least 300 eosinophils/μl in the previous 12 months.

Treatment compliance was strictly assessed at each clinical visit. Socio-demographic characteristics (age, sex, body mass index, smoking status, age at onset of asthma, sensitization to perennial aeroallergens) were included in the database as well as the presence of any comorbidities (nasal polyps, allergic rhinitis, GERD, nonallergic rhinitis with eosinophilia syndrome - NARES, obesity, bronchiectasis), which were objectively assessed according to standardized definitions and eventually confirmed by additional tests. This study used anonymous retrospective claims data, and as such, it did not require institutional review board review and approval or informed consent.

### 2.2. Measurements

The health records for each patient were recorded at baseline (T0), after 3 (T1), 6 (T3), nine (T6) and 12 months (T12) of treatment with MEPO. The following parameters were assessed: asthma control test (ACT) score, blood eosinophil count, forced expiratory volume in 1 second (FEV_1_), FEV_1_% of predicted and FEV_1_/FVC (forced vital capacity) ratio [15]. Spirometry was performed according to the ATS/ERS guidelines [16]. FEV_1_ and FVC were measured using a spirometer (Sensormedics, Milan, Italy). The best value of three consecutive maneuvers was expressed as the percentage of the normal value. After the baseline assessment, spirometry was repeated 15 minutes after administration of salbutamol (400 μg). Reversibility of airway obstruction was expressed in terms of percentage change from baseline FEV. Monthly intake (mg) of prednisone and exacerbations (per period of time, corrected per year and calculated as episodes requiring systemic corticosteroid treatment for at least 3 days, and/or emergency visit or hospitalization for acute asthma) were also included in the database for all time points

### 2.3. Evaluation of the response to mepolizumab

We selected five parameters which are crucial in the treatment of severe eosinophilic asthma and accordingly, patients were divided into two groups: responders and non-responders. Clinical relevant response was defined as: i) a 30% decrease in the exacerbation rate [17]; ii) an improvement in pulmonary function (FEV_1_ ≥ 200 mL) by analogy to the cut-offs used by the Global Lung Initiative [14]; iii) an 80% reduction of eosinophils in peripheral blood from baseline by analogy to the approval studies of mepolizumab [3-6]); iv) a change in ACT from baseline, whereby minimal clinically relevant difference was defined as an ACT score of three points [18]. We did not include OCS reduction as a clinical response parameter as not every single patient was on continuous OCS at T0. Fulfilling at least three of the three components of the primary outcome was considered a treatment success.

### 2.4. Statistical analysis

Statistical analysis was performed using GraphPad Prism v8.0 (Graphpad Software, Inc., La Jolla, CA). Categorical variables are stated as numbers (n) and percentages (%). Continuous variables are presented as mean ± SD (if normally distributed) or median and interquartile ranges (IQR) unless indicated otherwise. Fisher's exact test, Chi-squared test, two-sided independent t-test, Wilcoxon matched-pairs signed-rank test or Mann-Whitney-U-test were used as appropriate. The normality of data distribution was checked using the Shapiro-Wilk test. Logistic Regression models were created to determine the effects of comorbidities on the outcomes. For comparisons of more than two groups, one-way analysis of variance (ANOVA) or Kruskal-Wallis were used as appropriate. A p-value <0.05 was considered as statistically significant.

## 3. Results

### 3.1. Assessment of all patients

We analyzed the data from 31 patients (mean age 52.35 years; 58% females) with severe eosinophilic asthma and on treatment with MEPO. Patients’ socio-demographic characteristics, including smoking status and comorbid conditions, are displayed in Table 1.

**Table 1.**
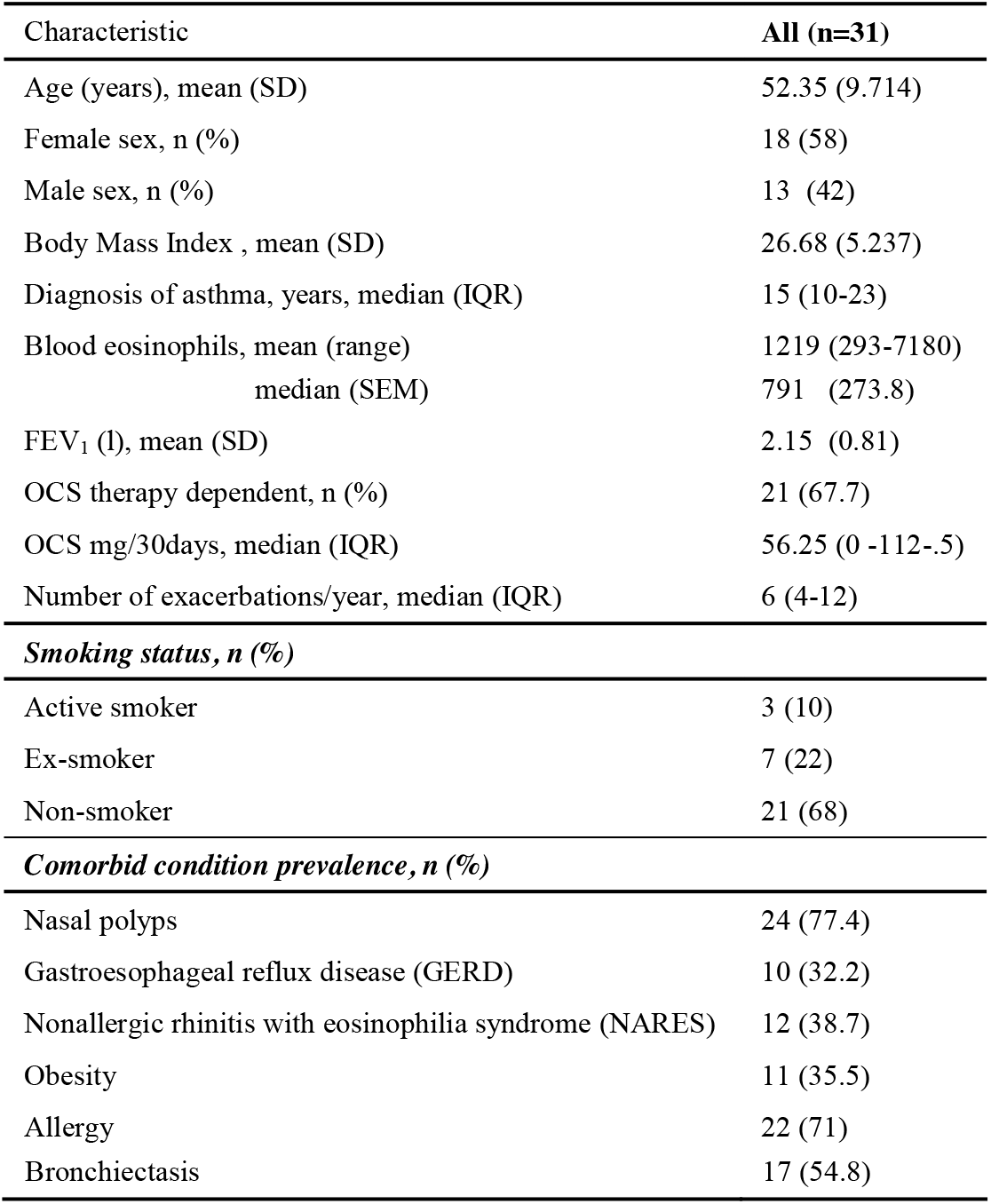
Demographic and baseline characteristics

The variables at baseline and 12 months (T12) are shown in Table 2.

**Table 2.** Summary of effectiveness outcomes

|  | **Before treatment** | **After treatment** | ***p* value** |
| --- | --- | --- | --- |
| **FEV<sub>1</sub> % of predicted, mean (SD)** | 73.68 (21.43) | 82.94 (21) | 0.0069 |
| **FEV<sub>1</sub> (L), mean (SD)** | 2.11 (0.748) | 2.33 (0.70) | 0.0224 |
| **FEV<sub>1</sub>/FVC, mean (SD)** | 69.48 (15.52) | 69.31 (11.23) | ns |
| **ACT, mean (SD)** | 13.65 (4.54) | 21.29 (4.49) | < 0.0001 |
| **Blood eosinophils, median (IQR)** | 791 (420 - 1300) | 80 (43-109) | < 0.0001 |
| **OCS therapy dependent, n (%)** | 21 (67.7) | 5 (16.1) | < 0.0001 |
| **OCS mg/30days, median (IQR)** | 56.25 (0 -112.5) | 0 (0-0) | 0.0012 |
| **Number of exacerbations/year, median (IQR)** | 6 (4-12) | 0 (0-1) | < 0.0001 |

The overall median blood eosinophil count decreased from 791 cells/ul (IQR 420-1300) at baseline to 80 cells/ul (IQR 43-109) at T12 (p>.0001). As shown in Figure 1A, the median decrease was already significant (p<0.0001) at the first time point (T1, 3 months) and was sustained at each consecutive time point.

**Figure 1.**
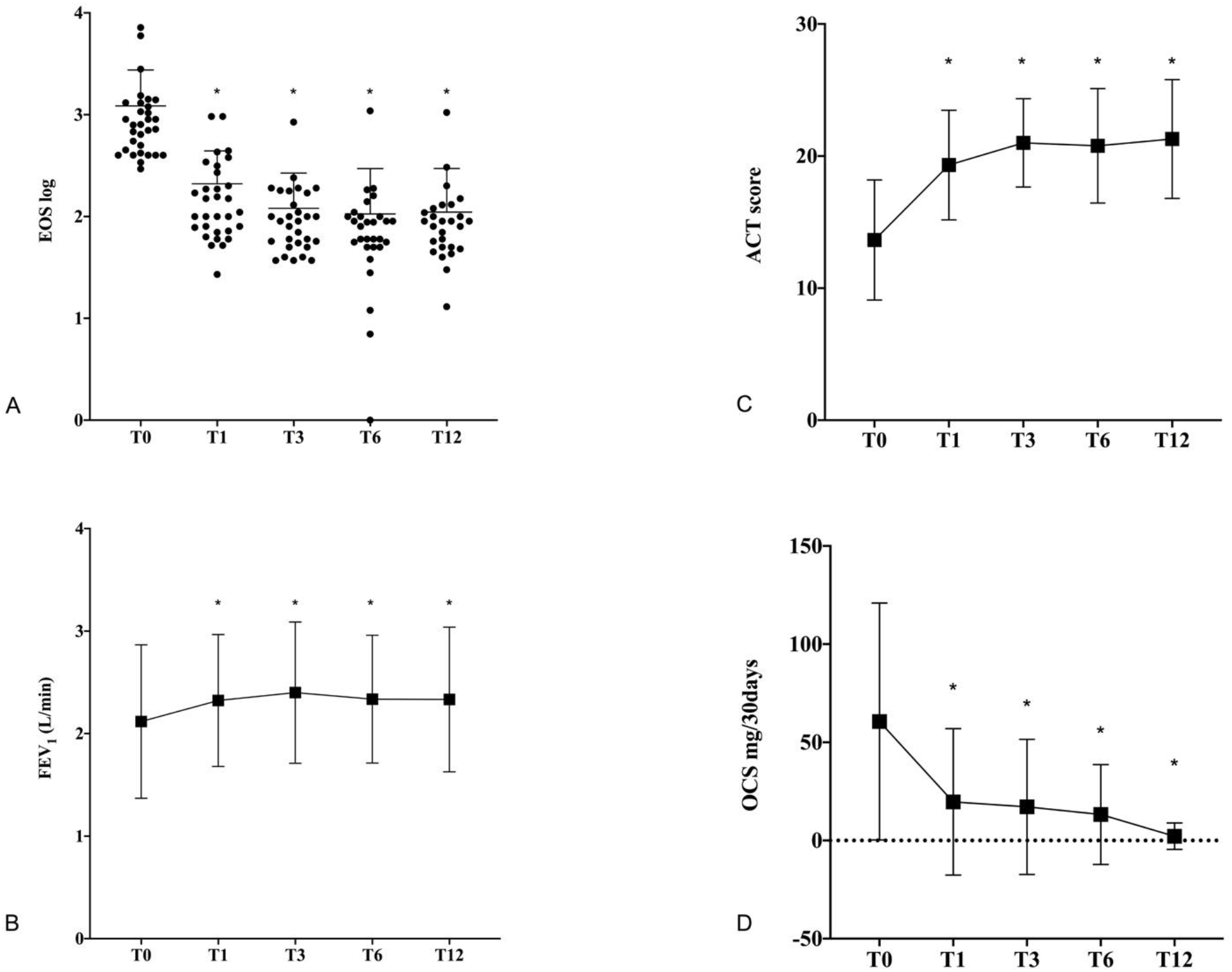
Comparison of clinical parameters from baseline (T0) to 12 months (T12). (A) Median blood eosinophil count; (B) Mean of FEV_1;_ (C) Mean of ACT score; (D) Median ofOCS mg/30days. * indicates p<0.05

A significant change in mean predicted FEV_1_ and FEV_1_% compared to baseline was observed at T12 (2.12±0.75 versus 2.33±0.7; p=0.0224) and (73.68±21.43 versus 82.94±21; p=0.0069), respectively. As shown in Figure 1B, mean FEV_1_ increase was already significant (p=0.0455) at T1 and was sustained at each consecutive time point. FEV_1_/FVC was significantly different from baseline only at T1 (79.19 ±16.38; p= 0.0036) and at T3 (82.71±16.24; p=0.0001), but not at T6 and T12 (data not shown).

There was a significant improvement in ACT after treatment with MEPO, with a mean of 13.65±4.54 points at baseline and 21.29±4.49 points at T12 (p>0.0001). As shown in Figure 1C, also for this outcome, the mean increase was already significant (p=0.0455) at T1 and was sustained at each consecutive time point.

At baseline, 67.7% of patients were on continuous OCS therapy with a median 30-days dose of 59.25 mg (IQR 0-112.5) of prednisone. Both OCS rates and dosage were significantly reduced at T12. Only 16.1% were still on OCS (p< 0.0001), with a lower median 30-day dose of 0 mg (IQR 0-0).

After one year of MEPO treatment, we observed a significant difference in the number of exacerbation/year (6, IQR 4-12 vs 0, IQR 0-1; p<.00001). All patients except one (96.77%) reduced their number of exacerbations by at least 70%. In particular, of the 17 patients (54.93%) who had more than 5 exacerbations in the year before therapy, 100% had no exacerbations at T12. Finally, no adverse effects were observed in our cohort.

### 3.2. Assessment of patients based on comorbidities

All patients had at least one comorbidity with a median (IQR) number of comorbidities of 3 (2-4). In detail: 3 patients (9.68%) had one comorbidity, 5 patients (16.13%) had two comorbidities, 13 patients (41.93%) had three comorbidities, 8 patients (25.81%) had four comorbidities, one patient had five comorbidities, and one patient had six comorbidities (data not shown). Females and males had a comparable number of comorbidities (3, IQR 3-4 vs 3, IQR1.5-4; p=0.248).

In order to evaluate the potential impact of comorbidities on treatment effect, a comparison of blood eosinophils count, ACT score, FEV_1_ values and OCS median dose among the six groups of patients with/without nasal polyps, GERD, NARES, obesity, bronchiectasis and allergy was performed. Table 3 shows that there was no significant difference at baseline between groups with/without comorbidities, with the only exception of ACT score between patients without and with GERD (12.52±5.47 versus 16±4.807, respectively; p=0.0443).

**Table 3.** Clinical parameters by comorbidities at baseline and at AT12-T0

| Table 3. Clinical parameters by comorbidities at baseline and at ΔT12-T0 | | | | | | |
| --- | --- | --- | --- | --- | --- | --- |
| Baseline | | | | ΔT12-T0 | | |
| Nasal polyps |  |  |  |  |  |  |
|  | Without (7) | With (24) | *p value* | Without (7) | With (24) | *p value* |
| FEV<sub>1</sub> (L) | 2.024 (1.002) | 2.18 (0.77) | ns | 0.284 (0.52) | 0.15 (0.508) | ns |
| ACT | 15.57 (5.028) | 13.08 (4.343) | ns | 5.286 (6.157) | 8.333 (4.556) | ns |
| OCS mg/30days | 112.5 (0 -112.5) | 16.88 (0-112.5) | ns | -112.5 (-112.5-0) | -11.25 (-11.25 -0) | ns |
| Exacerbations/year | 12 (4-12) | 6 (3.25-12) | ns | -11 (-12- -4) | -6 (-12- -2.25) | ns |
| Eosinophils | 1299 (500 - 1400) | 711( 400- 1063) | ns | -995 (-1200 - -500) | -640 (-927 - -347) | 0.0395 |
| GERD |  |  |  |  |  |  |
|  | Without (21) | With (10) | *p value* | Without (21) | With (10) | *p value* |
| FEV<sub>1</sub> (L) | 2.033 (0.782) | 2.398 (0.871) | ns | 0.259 (0.424) | 0.02 (0.64) | ns |
| ACT | 12.52 (5.47) | 16 (4.807) | 0.0443 | 8.048 (5.47) | 6.8 (4.022) | ns |
| OCS mg/30days | 56.25 (0 -112.5) | 67.5 (0 -112.5) | ns | -33.75 (-112.5 -0) | -56.25 (-112.5 -0) | ns |
| Exacerbations/year | 8 (4-12) | 6 (2.75-12) | ns | -7 (-12- -4) | -5.5 (-12- -2) | ns |
| Eosinophils | 800 (460-1135) | 755.5 (400-1764) | ns | -700 (-987.5 - -420) | -515.5 (-1569 - -386.8) | ns |
| Obesity |  |  |  |  |  |  |
|  | Without (20) | With (11) | *p value* | Without (20) | With (11) | *p value* |
| FEV<sub>1</sub> (L) | 2.295 (0.950) | 1.868 (0.515) | ns | 0.142 (0.355) | 0.09 (0.59) | ns |
| ACT | 14.85 (4.082) | 12.09 (5.224) | ns | 7.25 (4.529) | 8 (6.017) | ns |
| OCS mg/30days | 112.5 (2.813 - 112.5) | 22.5 (0-98.44) | ns | -112.5 (-112.5 – 2.813) | -21.5 (-111.5 -0) | ns |
| Exacerbations/year | 6 (4-12) | 8 (4-12) | ns | -6 (-12- -4) | -7 (-12- -3) | ns |
| Eosinophils | 711 (405 - 1200) | 1040( 550- 1418) | ns | -653.5 (-960.8 - -400) | -743 (-1110 - -368.0) | ns |
| Bronchiectasis |  |  |  |  |  |  |
|  | Without (14) | With (17) | *p value* | Without (14) | With (17) | *p value* |
| FEV<sub>1</sub> (L) | 2.133 (0.793) | 2,165 (0.858) | ns | 0.173 (0.49) | 0.189 (0.534) | ns |
| ACT | 13.5 (4.926) | 13.76 (4.352) | ns | 6.643 (5.583) | 8.471 (4.501) | ns |
| OCS mg/30days | 22.5 (0 -112.5) | 112.5 (0 -112.5) | ns | -11.25 (-112.5 -0) | -112.5 (-112.5 -0) | ns |
| Exacerbations/year | 4 (2.75-12) | 12 (5-12) | ns | -4 (-9- -2) | -11 (-12- -5) | ns |
| Eosinophils | 970 (437.5- 1449) | 720 (410 - 985) | ns | -685 (-1248 - -358.5) | -657 (-919 - -370) | ns |
| NARES |  |  |  |  |  |  |
|  | Without (19) | With (12) | *p value* | Without (19) | With (12) | *p value* |
| FEV<sub>1</sub> (L) | 2.197 (0.89) | 2.078 (0.712) | ns | 0.127 (0.469) | 0.269 (0.570) | ns |
| ACT | 13.79 (4.709) | 13.42 (4.461) | ns | 7.526 (5.621) | 7.833 (4.108) | ns |
| **OCS mg/30days** | 56.25 (0 -112.5) | 22.5 (0 -112.5) | ns | -56.25 ( -112.5 -0) | -22.5 (-112.5 -0) | ns |
| **Exacerbations/year** | 8 (4 -12) | 6 (3.25-12) | ns | -7(-12- -4) | -6 (-11.75- -2.5) | ns |
| **Eosinophils** | 898 (500 -1400) | 691 (400- 1235) | ns | -700 (-1110 - -368.0) | -653.5 (-983.8 - -345.8) | ns |
| **Allergy** |  |  |  |  |  |  |
|  | **Without (9)** | **With (22)** | **p value** | **Without (9)** | **With (22)** | **p value** |
| **FEV1 (L)** | 2.421 (0.675) | 1.995 (0.7558) | ns | 0.136 (0.571) | 0.2 (0.49) | ns |
| **ACT** | 14 (4.5) | 13.5 (4.657) | ns | 4.778 (4.764) | 8.818 (4.727) | 0.048 |
| **OCS mg/30days** | 56.25 (0 -112.5) | 39.38 (0 -112.5) | ns | -33.75 ( -112.5 -0) | -39.38 (-112.5 -0) | ns |
| **Exacerbations/year** | 12 (2.5 -12) | 6 (4-12) | ns | -8 (-11.5- -1.5) | -6 (-12- -4) | ns |
| **Eosinophils** | 702 (410 -1364) | 795.5 (437.5- 1225) | ns | -500 (-979-5 - -333.5) | -721.5 (-1024 - -398.3) | ns |

ACT are expressed ad mean (SD), OCS mg/30days as median (IQR), Number of exacerbations/year as median (IQR), Eosinophils as median (IQR), FEV<sub>1</sub> as mean (SD).

We did not find any significant difference for ΔT12-T0 in any of the analyzed clinical parameters among patients with and without comorbidities, with the only exceptions of patients without nasal polyps, who showed a greater blood eosinophil reduction than those with nasal polyps (−995, IQR - 1200 - −500 vs −640, IQR −927 - −347; p=0.0395), and patients with allergy, who showed a greater ACT score than those without allergy (8.818±4.727 vs 4.778±4.764; p=0.048) (Table 3).

The association between the median number of observed comorbidities and the ΔT12-T0 of ACT mean scores (1-2 comorbidities: 4.625±5.878; 3 comorbidities: 8.615±5.06; ≥ 4 comorbidities: 8.8±3.46; p=0.1412), of FEV_1_ mean values (1-2 comorbidities: 0.22±0.304, 3 comorbidities: 0.1662±0.537, ≥ 4 comorbidities: 0.173±0.628; p=0.9719), OCS median dose (1-2 comorbidities: - 112.5, IQR-140.6 - −19.69, 3 comorbidities: −112.5, IQR −112.5 - −90, ≥ 4 comorbidities: −84.38, IQR −112.5 - −30.94; p=0.7741) and median blood eosinophilia (1-2 comorbidities: −645.5, IQR - 109.5 - −380, 3 comorbidities: −657, IQR −1087 - −380, ≥ 4 comorbidities: −696.5, IQR −1013 - −375; p=0.9933) was not significant.

### 3.3. Clinical response

According to our clinical response parameters, 83.87% (26) of patients were classified as responsive to MEPO treatment. A substantial depletion of the blood eosinophils (less than 80% from baseline) was found in 87.1% of patients, improvement in lung function (FEV_1_ > 200 mL) was seen in 17 patients (54.84%), 3-point improvement in ACT from baseline was recorded in 25 patients (80.65%) and a 30% reduction of exacerbation rates was seen in 30 patients (96.77%). Moreover, the majority of patients (38.71%) met 3/4 parameters after 12 months, as shown in Table 4.

**Table 4.** Response to treatment with mepolizumab

| **Table 4. Response to treatment with mepolizumab** |  |  |  |
| --- | --- | --- | --- |
|  |  | **N** | **%** |
| **Overall response** |  | 26 | 83.87 |
| **Outcome summary** |  |  |  |
| **(number of fulfilled parameters)** |  |  |  |
|  | 4/4 | 12 | 38.71 |
|  | 3/4 | 14 | 45.16 |
|  | 2/4 | 4 | 12.9 |
|  | 1/4 | 1 | 3.22 |
|  | 0/4 | 0 | 0 |
| **With Nasal polyps** |  | 21 | 87.5 |
| **With GERD** |  | 8 | 72.72 |
| **With NARES** |  | 9 | 75 |
| **With Obesity** |  | 9 | 87.5 |
| **With Allergy** |  | 18 | 81.82 |
| **With Bronchiectasie** |  | 14 | 82.35 |

The characteristics of the five non-responding patients are summarized in Table 5.

**Table 5.** Characteristics of non-responder patients

| Patient | Age | Comorbidity | Missed Outcome |
| --- | --- | --- | --- |
| A | 70 | Bronchiectasie | FEV<sub>1</sub>↑, ACT↑ |
| B | 46 | Nasal polyps, GERD, Obesity | FEV<sub>1</sub>↑, ACT↑, Blood eosinophils↓ |
| C | 69 | GERD, NARES, Allergy | FEV<sub>1</sub>↑, ACT↑ |
| D | 55 | Nasal polyps, NARES, Allergy, Bronchiectasie | FEV<sub>1</sub>↑, ACT↑ |
| E | 41 | Nasal polyps, GERD, NARES, Allergy | FEV<sub>1</sub>↑, Blood eosinophils↓ |

### 3.4. Predictive factors

In order to identify potential predictive factors of MEPO response, we analyzed if every single comorbidity, and smoking status, gender (female), age ≥65 years-old, BMI ≥ 25kg/m^2^ and blood eosinophil count ≥ 500/mm^3^ of these 31 patients were associated with allocation to a specific treatment response group (responders or non-responders). As shown in Figure 2, each of the analyzed variables achieved a significant value of p>0.05 in the univariate model; thus none of them influenced allocation to a specific treatment response group.

**Figure 2.**
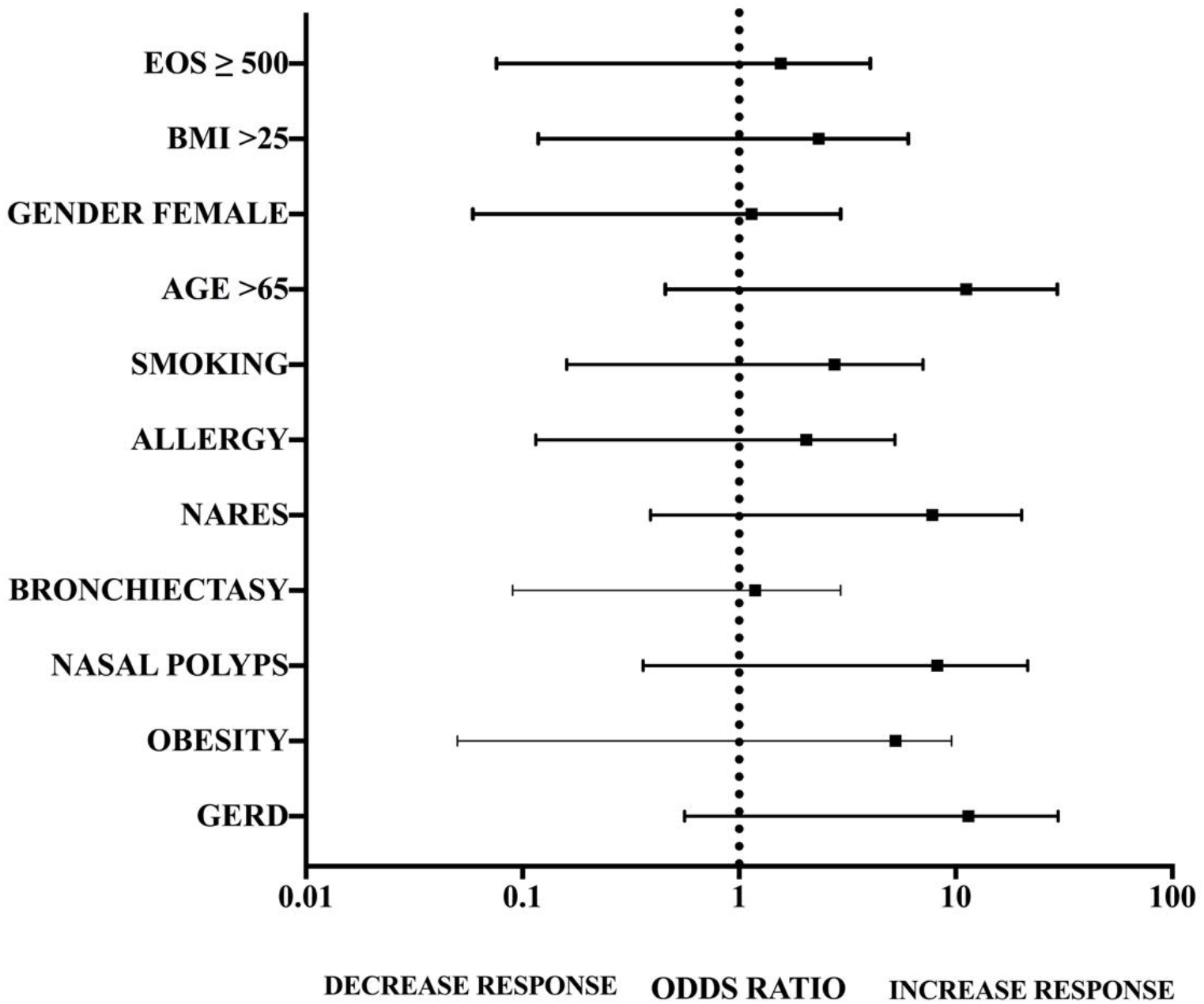
Analysis of potential predictors of treatment outcome.

## 4. Discussion

To the best of our knowledge, this is the first study assessing not only the efficacy of mepolizumab in patients with severe eosinophilic asthma complicated by the presence of one or more comorbidities but also whether these affected the treatment outcome or not.

Our first analysis led us to conclude that the treatment with mepolizumab for one year substantially improved all the analyzed clinical parameters. Mepolizumab resulted in a significant reduction in asthma exacerbations, use and dose of OCS, blood eosinophilia and a concomitant improvement in pulmonary function and asthma symptom control in all patients.

The overall response rate was of 83.87%. In particular, blood eosinophil count decreased by 89.89%, a 3-point improvement in ACT from baseline was recorded in 80.65% of patients and exacerbation rates were virtually zeroed, as 96.77% of patients had a reduction in the number of exacerbations by minimum 30% and at least 70% during the year of treatment with mepolizumab. Also, a sharp reduction in the use of OCS was recorded in our cohort, as 84% of patients discontinued the OCS at follow-up, a percentage higher than so far reported in other studies [19-23].

In our cohort, FEV_1_ values increased only by 9% and the FEV_1_/FVC difference between follow-up and baseline was statistically significant at 3 months but not at 12 months; these data seem to align with those of other studies [21]. The FEV_1_/FVC result could be explained as a concomitant increase of airway caliber and reduction in the residual volume (RV), which usually takes place in response to asthma treatment and improves both FEV_1_ and FVC [24].

Overall, our results are comparable with those attained in both randomized and real-life analyses [3-6,20-23,25,26]. Also, it is important to underline that our data not only confirm the efficacy of MEPO, but also highlight the rapidity of the therapeutic effect. In our study, a significant improvement in FEV_1_ and blood eosinophil count was already evident after 3 months and was sustained for 12 months. The quick beneficial effect of MEPO was in accordance with the reported patient’s ACT score, which also significantly improved within the first 3 months. An equally rapid response, even within the first month of treatment, has been highlighted in other real-life studies [19,20,22,23,25].

In our cohort, only 5 patients did not exhibit, according to our criteria, an effective response to treatment These patients had an average age of 56.2 years old (minimum 50, maximum 70) and had distinctive comorbidities or combinations of comorbidities without a recurrent pattern. The outcome that was mostly not achieved among these five patients was the increase of 200 mL in FEV_1_, followed by the 3-point increase in the ACT score, and in only two patients the 80% decrease in the level of eosinophils in the blood. However, our clinical response cut-off was particularly stringent and, in general, the clinical conditions of these patients were ameliorated by MEPO therapy.

Our second analysis questioned whether patients with specific comorbidities achieved different results in treatment outcomes. Among patients with or without a comorbidity, we did not find any statistically significant difference. The only exceptions were patients without nasal polyps, who showed a more significant reduction in blood eosinophilia than patients with nasal polyps (p=0.0395), and patients with allergy, who showed a more considerable improvement in their ACT score than those without allergy (p=0.048). Not even the number of comorbidities influenced treatment with MEPO, as no difference in achieving the therapeutic success was found among patients having 1-2 comorbidities, 3 comorbidities or more than 4 comorbidities.

These data are particularly useful to assess the role of mepolizumab better as we provide an insight into real-life characteristics of all eligible patients. Our 31 patients had a median number of 3 comorbidities, a situation that differed largely from that of RCTs, in which patients do not present any concomitant disease. In this regard, a recent real-life study by Bagnasco and colleagues compared the characteristics at baseline of its cohort with those of patients enrolled in MEPO RCTs [27]. Their results underline how real-life patients were characterized by a greater age, a worse lung function, a higher level of eosinophilia and a higher dosage of OCS compared to RCT patients [27]. Comparing the baseline characteristics of our cohort with the cohort of the study of Bagnasco and colleagues, it is possible to observe an even higher level of eosinophilia at baseline (653 ± 381 vs 1219 ± 1585 respectively; p = 0.0034), a similar baseline level of FEV_1_% and a greater annual recurrence of exacerbations (3 ± 1.8 vs 7.58 ± 4.178; p < 0.0001).

Taken together, these data suggest that mepolizumab is capable of exerting its beneficial action in patients with severe eosinophilic asthma despite the presence of one or more comorbidities.

To date, only two studies have assessed the effectiveness of MEPO in patients with comorbidities, and both corroborate our data [28-29]. The first study has evaluated MEPO outcomes after 12 months of treatment in four severe uncontrolled asthmatic patients with bronchiectasis [28]. Results revealed a significant increment in ACT and lung function, a reduction in the number of exacerbations/year and a reduction of blood eosinophilia [28]. The second study has found a correlation between the presence of eosinophilic chronic rhinosinusitis (ECRS) and therapeutic response in patients with severe eosinophilic asthma [29]. In particular, Numata and colleagues identified in 28 patients that ECRS was a predictive factor of the response to mepolizumab as patients with eosinophilic chronic rhinosinusitis showed significantly improved systemic corticosteroid-sparing effects, lung function and symptoms compared to patients without the comorbidity [29].

In order to extend our analysis, we probed if single comorbidities influenced allocation to the responder or non-responder group. Neither the comorbidities nor other characteristics of patients at baseline (i.e. sex, BMI, age, smoking habits, baseline eosinophil count) affected the success or failure of MEPO therapy. Other studies evaluated some socio-demographic factors (e.g. allergy, BMI, eosinophils and lung function at baseline, age, sex and smoking habits) and two of them identified potential predictive factors of MEPO response [25,30,31]. A supervised cluster analysis with a recursive partitioning approach applied to the Dose Ranging Efficacy And safety with Mepolizumab (DREAM) data identified BMI as a predictor.^30^ However, the data on this topic are rather controversial, and there is no general agreement as to the role of BMI as a predictive factor of outcome [25,30].

Finally, like other real-life studies, our data do not indicate the number of eosinophils as a predictor of clinical efficacy, suggested by some other Authors as a useful biomarker for the selection of patients who are more likely to benefit from treatment with MEPO [21,25,30,32,33].

There are several limitations to the present study. One limitation is that it is a single-center, retrospective study. However, the alignment of our results with those from the literature makes data more robust. Secondly, the conclusions about predictive factors could be limited due to the small number of patients included in the study. Thirdly, no established criteria for treatment response have been validated yet; therefore our criteria could be classified as subjective.

## 5. Conclusions

Asthma is a heterogeneous disease, including very different clinical conditions. Therefore, the systematic investigation of flawless biomarkers or composite indexes which could help clinicians identify patients predisposed to specific therapeutic strategies is still an unmet need.

These findings, while preliminary, suggest that treatment with MEPO is effective in the clinical practice in patients with severe eosinophilic asthma complicated by one or more comorbidities. However, as we were not able to establish a predictive outcome factor, further larger studies, which take these variables into account, will need to be undertaken.

## Data Availability

The authors confirm that the data supporting the findings of this study are available within the article [and/or] its supplementary materials.

## Acknowledgements

Editorial assistance for the manuscript was provided by sciencED and supported by GSK.

## Abbreviations

ACT: asthma control test
EA: eosinophilic asthma
ECRS: eosinophilic chronic rhinosinusitis
FEV_1_: forced expiratory volume in 1 second
FVC: forced vital capacity
GERD: gastro-esophageal reflux disease
MEPO: mepolizumab
NARES: nonallergic rhinitis with eosinophilia syndrome
OCS: oral corticosteroid
RCTs: randomized controlled trials

